# Momentary Brain Imaging Assessment of Neuroplasticity in Psychiatric Illness: Protocol for Systematic Review

**DOI:** 10.1101/2025.02.19.25322078

**Authors:** A. Brown, S. Upadhyay, E. C. Lloyd

**Affiliations:** Columbia University Teachers College; Columbia University Irving Medical Center

**Author notes:** Joint first author (these authors contributed equally to the manuscript) (A. Brown, S. Upadhyay). Corresponding author information: Amanda Brown, Address: 525 W 120th St, New York, NY 10027, Shivani Upadhyay, Address: 319 W 101st St, New York, NY 10025, E. Caitlin Lloyd.

**Keywords:** Neuroplasticity, brain plasticity, neuroimaging, psychiatric disorder, biomarkers

## Abstract

**Background:** Neuroplasticity describes the brain’s ability to adapt in response to alterations in the environment, and is fundamental to learning. Reduced neuroplasticity has been proposed to underlie several psychiatric symptoms and disorders. Advances in neuroimaging have provided new methods for examining or indexing the potential for neuroplastic changes in-vivo. The proposed systematic review and meta-analysis will synthesize research utilizing neuroimaging assessments to compare the potential for rapid brain changes between individuals with commonly-studied psychiatric disorders relative to healthy control peers (HC).

**Methods:** This systematic review will include studies comparing the potential for neuroplastic changes between individuals with commonly-studied psychiatric disorders (mood, anxiety, compulsive, trauma-related, eating, and schizophrenia and other psychotic disorders). Included studies will assess neuroplasticity using established or putative neuroimaging biomarkers. Longitudinal studies, studies using non-neuroimaging methods to assess neuroplastic potential, and animal studies will be excluded. PubMed, Web of Science, Embase, and PsycINFO will be searched using predefined terms. Two independent reviewers will screen titles, abstracts, and full texts using Rayyan, with conflicts resolved by a third reviewer. Data including study and participant characteristics will be extracted. Summary statistics will be combined and analyzed using random-effects meta-analyses to estimate the average difference in neuroplasticity between groups. In the event of heterogeneity, subgroup analyses and meta-regression will explore potential moderators of the between-group difference. The extent to which publication bias is likely to impact the findings of the review will be assessed using Egger’s test.

**Discussion:** This review will summarize alterations in neuroplasticity, as indicated by momentary neuroimaging assessments, among individuals with major psychiatric disorders. As research increasingly links psychiatric conditions to neuroplasticity, this review will offer a valuable resource for understanding how neuroplasticity can be measured in-vivo to examine mechanisms of psychiatric illness.

**Systematic review registration:** This review is registered in PROSPERO (registration number: CRD42025630626)

## Background

Neuroplasticity refers to the brain’s ability to reorganize itself in response to experience through structural and functional changes that modify neural interactions. Neuroplasticity allows the creation of new neural pathways and the strengthening or weakening of existing ones, to support learning and behavioral adaptation. Reductions in neuroplasticity have been suggested to contribute to impaired memory, learning and flexibility, and consequent inflexibility in psychiatric disorders (e.g., (1–3)). With advances in non-invasive neuroimaging techniques it is increasingly possible to characterize neuroplasticity in-vivo using momentary assessments (4–6). The proposed review will characterize the use of these assessments in examining neuroplasticity in psychiatric illness, and summarize observed patient and healthy control (HC) differences.

Neuroplasticity encompasses a broad range of changes to the nervous system, including the proliferation of new-born neurons (neurogenesis), structural changes to the axons and dendrites of neurons (e.g., including axonal and dendritic arborisation and pruning) that affect synaptic density, and modification of synapse strength. While neuroplasticity is much higher during development, changes to brain structure and function in response to experience occur throughout life, allowing individuals to learn about and adapt to their environments (7–13).

While neuroplasticity is necessary for adaptive behavior, disturbances relating to, and particularly a loss of, neuroplasticity are hypothesized to contribute to the onset and maintenance of various psychiatric diagnoses. Anxiety disorders and major depression are proposed to result from decreased plasticity (particularly within frontolimbic circuits), leading to inflexible patterns of thinking (e.g., worry, rumination), and deficits in memory and learning (e.g., extinction and safety learning) that serve to maintain psychiatric symptoms (1,2). Similarly, reductions in neuroplasticity have been proposed to contribute to difficulty modifying eating-related symptoms in anorexia nervosa (14), and cognitive deficits in schizophrenia (15). Animal models and postmortem studies have implicated reduced neurogenesis and synaptic plasticity in anxious and depressive behavior (16,17), and suggest impaired learning as a mechanism of effect (3,18). Furthermore, pharmacological interventions that enhance neuroplasticity, for example ketamine, show efficacy in the treatment of depression and anxiety disorders (19–22).

While assessments of neuroplasticity in humans were previously limited to post-mortem investigations, developments in neuroimaging methodologies increasingly allow experience-dependent changes, or the potential for these changes, to be assessed in the moment, in-vivo. Structural MRIs and Diffusion Tensor Imaging (DTI) have been used to identify experience-dependent changes in brain volume and connectivity, respectively. For example, studies have identified within-session changes in gray and white matter volumes that occur with new learning (23,24). The examination of learning-related changes in brain function has been probed through combining stimulus presentation with functional imaging. In these paradigms, neuroplasticity is indexed by changes in neural responses following exposure to a stimulus (e.g., a tone (25,26))). A popular paradigm for examining neuroplasticity involves the pairing of motor cortex stimulation (using transcranial magnetic stimulation) with median nerve stimulation, which is known as paired associative stimulation (4). In this paradigm the amplitude of motor-evoked potentials increases, and the amount of stimulation needed to evoke a motor response reduces, as a result of the repeated afferent signals from the somatosensory system (i.e., due to peripheral nerve stimulation) that co-occur with motor cortex activity. These effects are reversed (i.e., motor response reduces) when somatic stimulation reaches the motor cortex after motor cortex stimulation. Changes in behavioral response are thought to result from changes in synaptic strength (enhanced when postsynaptic activity is preceded by presynaptic activity), and have been localized and quantified in the brain using electroencephalography (EEG; (27)). Developments in this method have included studying paired associative stimulation of non-motor regions, and have similarly quantified the effects on brain activation (or plasticity) using EEG (28,29).

Other studies have examined individual or group differences in neuroplasticity by using putative neuroimaging markers of neuroplastic status or potential. Several such markers are related to the excitation-to-inhibition (E/I) ratio. The opening of critical periods of neuroplasticity during development depends on reducing the E/I (30). This change in E/I is driven by the maturation of inhibitory circuits that suppresses spontaneous activation thereby improving the signal-to-noise ratio of stimulus-evoked activity (31). In contrast, neuroplasticity in adulthood may actually require a reduction in inhibitory tone (32). Both functional MRI and EEG assessments have been validated to quantify inhibitory activity or E/I (for recent overview, see (33)) and thus used to examine developmental changes in plasticity potential.

Though neuroimaging assessments have the potential to provide insights into whether individuals with psychiatric illness have differential neuroplasticity, the extent to which they have been used to test mechanistic hypotheses, and the overall pattern of findings, is unclear. While individual studies have reported the use of putative markers of plasticity or plasticity potential to compare patients with psychiatric illness and HC (e.g., (34–40)), a comprehensive overview of this literature is lacking. Such an overview is critical to understand the methods available to assess neuroplasticity as well as the evidence concerning alterations present in psychiatric illness. The proposed systematic review will fill this gap in the literature to provide a summary of the science surrounding neuroplasticity in psychiatric illness that will guide future research and practice. The review will address the question of whether momentary neuroimaging markers of neuroplasticity differ between patients with a DSM-5 psychiatric disorder compared to individuals without a DSM-5 psychiatric disorder. The findings will offer insights into whether neuroplasticity might be leveraged to improve cognitive and behavioral dysfunctions, offering a foundation for the development of novel interventions across psychiatric conditions.

## Methods/Design

### Eligibility criteria

We will include all studies that assess neuroplasticity using neuroimaging techniques in individuals with and without commonly studied DSM-5 psychiatric disorders (Schizophrenia Spectrum and Other Psychotic Disorders, Bipolar and Related Disorders, Depressive Disorders, Anxiety Disorders, Obsessive-Compulsive and Related Disorders, Trauma and Stressor-Related Disorders, and Feeding and Eating Disorders) (41). Only human studies assessing putative or established biomarkers of brain plasticity, or neuroplastic potential, will be included. Studies examining immediate brain changes in response to stimuli in the same neuroimaging session (i.e., providing a momentary assessment of plasticity) are included. Studies examining changes over a longer timescale (i.e., days, weeks, years) are not, given these are likely assessing a different facet of neuroplasticity compared to momentary measures. We will exclude developmental studies, longitudinal studies, animal studies, and studies that use non-neuroimaging methods to assess neuroplasticity, as these are beyond the scope of our research question.

### Population

Our study population will include individuals of any age diagnosed with the following DSM-5 psychiatric disorder: Schizophrenia Spectrum and Other Psychotic Disorders, Bipolar and Related Disorders, Depressive Disorders, Anxiety Disorders, Obsessive-Compulsive and Related Disorders, Trauma and Stressor Related Disorders, Feeding and Eating Disorders. These constitute commonly-studied psychiatric disorders, ensuring our results will be broadly applicable and relevant to a wide range of future research (41). Furthermore, these illnesses share a genetic liability, suggesting it is helpful to consider shared brain mechanisms of illness across these disorders (42).Comparisons will be made with individuals without a diagnosis of a DSM-5 psychiatric disorder. Studies involving individuals with neurological disorders will be excluded to maintain focus on psychiatric populations.

### Index comparator

The primary comparison will be between individuals with a diagnosis of a DSM-5 psychiatric disorder (restricted to included diagnoses) and individuals without a diagnosis of a DSM-5 psychiatric disorder (healthy controls).

### Outcomes

The primary outcome measures are neuroimaging assessments of neuroplasticity. These measures should provide single-point-in-time assessments of neuroplastic potential. That is, studies examining changes in brain structure and function over periods that are not constrained to a single study session are excluded. Included in-vivo assessments of neuroplastic potential may use any neuroimaging method, such as magnetic resonance imaging (MRI), functional MRI (fMRI), transcranial magnetic stimulation (TMS), positron emission tomography (PET), diffusion tensor imaging (DTI), magnetoencephalography (MEG), electroencephalography (EEG), voxel-based morphometry (VBM), and mean diffusivity from diffusion imaging.

### Data sources

We will identify eligible studies by searching the PubMed, Web of Science, Embase, and PsycINFO databases for studies that were published (online or in-print) prior to December 2024.

### Search strategy

The detailed search strategy is described in Appendix 1, and was developed following extensive preliminary searches and consultation with an information specialist at Teachers College library. The following search terms were used: “neuroplasticity, brain plasticity, MRI, neuroimaging, brain, psychiatric illness, mental illness”. For the full search strategy see the appendix.

### Study selection and screening process

Titles identified from the primary literature search will be screened for eligibility based on title, abstract, and full text. The freely-available software Rayyan (43) will be used to screen studies and record decisions about eligibility. Study screening (at each level) will be performed by two independent reviewers. Titles rejected by both reviewers will be excluded, and the remaining studies will undergo abstract screening. Abstracts rejected by both reviewers will similarly be excluded; each full-text article passing abstract screening will be screened by two investigators for inclusion in the final dataset. Conflicts during full-text screening will be resolved by a third reviewer. Kappa scores (44) indexing the level of agreement reached between the reviewers at each stage of screening will be reported, as will the rationale for the inclusion and exclusion of studies at each screening stage. To ensure the capture of all relevant studies, we will review the references of all included articles, and review the studies that cite included studies in order to identify additional studies meeting inclusion criteria that may have been missed by the database searches.

### Data extraction and management

Pertinent data from each study (see below) will be extracted and stored in Rayyan (43).

### Data items

Extracted data will include study characteristics (title, authors, year of publication, country, journal, study design, sample size, and inclusion/exclusion criteria); and participant characteristics (age, sex, ethnicity, psychiatric illness, duration of illness, illness severity). We will record the assessment method for the disorder, the sampling/recruitment method of participants, the measure of neuroplasticity, details of how this measure was collected (MRI/EEG protocols and procedures for data collection and preprocessing), details of the analysis conducted, and measurement validation details. Means and standard deviations of the measure(s) of neuroplasticity for each group in each study will be extracted to calculate effect sizes. We will also extract information about associations between neuroplasticity assessments and psychiatric symptoms. Data will be extracted by two members of the research team and discrepancies resolved by a third member. The agreement between reviewers (Cohen’s kappa) (44) will be reported. Study authors will be contacted in the event data items are missing.

### Assessment of quality and risk of bias in included studies

All included studies will be thoroughly evaluated in regard to their different biases (name types of biases, selection etc.) according to the study design. We will assess the selected studies using the Downs and Black instrument (45). We will modify this instrument to remove questions relevant to intervention studies. Two reviewers will conduct the risk of bias assessment, independently. The agreement between reviewers will be reported for risk of bias assessment.

### Data synthesis and analysis

The characteristics of participants in included studies will be extracted and summarized. Random-effects meta-analytic models (46) describing the (weighted) average difference between neuroplasticity of patient and control samples will be estimated using R statistical software (47). Random effects models, rather than fixed effects, will be used as we anticipate substantial variation between the studies (46). All individual study effect sizes quantifying patient-HC differences will be converted into the standardized mean difference, represented with Hedge’s *g*, prior to conducting the meta-analysis. Hedge’s g provides a corrected estimate for reducing the upward bias that typically occurs in small sample sizes (48).

Heterogeneity of effect sizes will be indexed by calculating *Q* and *I*^2^ statistics. The *Q* statistic indicates whether differences between the effect sizes of the included studies are larger than expected due to chance (49). The *I*^2^ statistic indexes the extent of heterogeneity amongst effect sizes (50). An *I*^2^ of 50% or higher indicates meaningful variance between effect sizes; if observed, subgroup analyses will be conducted for studies categorized by disorder and measure of plasticity. If possible we will conduct meta-regression analyses (51) to determine whether psychiatric disorder, neuroplasticity measure, and sample characteristics (age, % males) moderate the magnitude of difference between patient and control groups.

We will use the Egger test to examine the potential for publication bias to affect results. In this approach the standardized effect sizes are regressed onto their precisions (standard errors). A significant intercept provides evidence of publication bias. To visualize the potential bias we will use the funnel plot (52) in which study estimates are plotted (around the meta-analysis mean estimate) against their precision. The trim and fill procedure imputes effect sizes that might be missing from the funnel plot (as indicated by asymmetry in the plot), to provide an adjusted effect size estimate that is less affected by publication bias (53). This procedure will be undertaken should publication bias be indicated.

If there are insufficient studies to conduct a meta-analysis (n < 5), we will conduct a qualitative synthesis only. Similarly, if sub-group statistical analysis is not possible due to the low number of studies in each category (n < 5), we will provide a narrative synthesis of patient-control differences in which studies are grouped by psychiatric illness of focus and measure of neuroplasticity.

A qualitative description of associations between psychiatric symptoms and neuroplasticity measures will be provided. This will be organized by psychiatric disorder.

A sensitivity analysis (54) will be conducted for the main meta-analysis comparison of patients and HC, including only studies that have been rated as low risk of bias. A separate sensitivity analysis will include only studies using validated measures of neuroplasticity.

The GRADE framework will be used to examine the quality of the overall body of evidence (55). The GRADE rating will inform the extent to which confidence should be placed in review conclusions.

## Discussion

This systematic review will identify methods that have been used to examine neuroplasticity among individuals with psychiatric illness and will summarize findings related to alterations in neuroplasticity among these individuals relative to HC. While a growing number of theoretical models of psychiatric illness implicate alterations in neuroplasticity in disorder symptoms (e.g., (1,2,14,33,56,57)), to our knowledge there has been no summary of outcomes of the studies testing model hypotheses. Furthermore the range of assessments that may be used to index neuroplasticity is unclear. The proposed review will thus provide an important resource for investigators interested in building upon existing knowledge to better understand disturbances in neuroplasticity in psychiatric disorders.

While narrative reviews of neuroplasticity assessment tools exist (e.g., (33,57)), a systematic review of studies using these techniques does not, and there has been no systematic summary of neuroplasticity research among psychiatric populations. The proposed review will involve the searching of multiple databases and reference searching to ensure capturing all relevant literature. The focus on multiple psychiatric disorders will promote the inclusion of a greater number of methods to assess neuroplasticity, as well as for identifying shared mechanisms of illness across psychiatric disorders. To ensure the inclusion of multiple diagnostic groups does not complicate the unbiased interpretation of results, where indicated we will consider the findings within distinct subgroups of patients or neuroplasticity assessments.

The review includes only momentary assessments of neuroplasticity. That is, studies examining changes in the brain over longer time-periods to index the plasticity of the brain are not included. Studies examining longitudinal changes over longer timescales are likely measuring a different quantity to momentary assessments, which may be better characterized as physiological measures of neuroplastic potential. Furthermore, group differences in longitudinal studies are likely at greater risk of influence by factors of no interest (given the range of events that may occur between initial and follow up scans) potentially complicating the interpretation of differences between patient and HC groups.

The strengths of this review include its broad search terms and inclusion criteria that will allow for identifying relevant articles, the carefully designed screening and bias assessment procedures, and the extraction of data by two members of the research team. This will ensure the synthesis of a wide range of information that is relevant to the research question, and that conclusions are based on high-quality data. The use of the GRADE criteria (55) will allow for providing information to the reader about the extent to which findings from this review are based on high-quality evidence, ensuring findings direct the research community to an appropriate extent.

The potential challenges include failure to identify relevant articles (e.g., due to search terms not being sensitive). This risk is mitigated by consulting with an expert librarian to refine the search strategy and using a range of databases in which the search will be implemented. Other challenges include the potential diversity of included studies (diverse in population, assessment, psychiatric illness) making it difficult to combine and compare them.For this reason we will quantify heterogeneity between studies and where appropriate meta-analyze, or qualitatively synthesise, them separately. A further challenge is the possibility of publication bias affecting inferences concerning patient-HC differences in neuroplasticity. Steps to identify and mitigate this issue (e.g., Egger test (52), Trim and Fill procedure) are however built into this protocol.

The major limitation of our review is that it includes cross-sectional evidence. Consequently, the findings will not be able to offer firm evidence as to whether or not neuroplasticity alterations constitute risk or maintaining factors for psychiatric illness. Nonetheless, currently it is unclear whether there are differences in neuroplasticity in psychiatric groups, which is necessary to establish to determine the validity of theories positing neuroplasticity alterations as mechanisms of illness. The findings may therefore provide a rationale for conducting longitudinal or experimental studies that require more resources, and which are better able to provide information about causality. Another limitation is that studies may use poorly validated assessments of neuroplasticity, rendering results invalid. The method of neuroplasticity assessment and validation of this metric will be recorded, allowing for examination of the extent to which metrics index neuroplastic potential of the brain. Furthermore, a sensitivity analysis in which studies using unvalidated assessments of neuroplasticity are excluded, will be conducted.

Overall, the review will provide an overview of the evidence for disturbances in neuroplasticity in psychiatric illness, and an overview of the research tools that have been used to address questions surrounding these disturbances. Outcomes will identify gaps in knowledge and the need for novel momentary assessments of neuroplastic potential. Ultimately, the findings will guide mechanism-based research in psychiatry, and the development of treatments able to target mechanisms of illness.

## Supporting information

PRISMA-P Checklist

Search Strategy

## Data Availability

All data produced in the present study are available upon reasonable request to the authors.

## List of abbreviations

DTI: Diffusion Tensor Imaging
EEG: Electroencephalography
E/I: Excitation-to-inhibition
fMRI: Functional MRI
HC: Healthy control
MEG: Magnetoencephalography
MRI: Magnetic resonance imaging
PET: Positron emission tomography
TMS: Transcranial magnetic stimulation
VBM: Voxel-based morphometry

## Declarations

### 1. Ethics Approval and Consent to Participate

Not applicable.

### 2. Consent for Publication

Not applicable.

### 3. Availability of Data and Material

All data generated or analyzed during this study will be included in the published systematic review and meta-analysis, and available as supplementary materials or upon reasonable request.

### 6. Authors’ Contributions

AB and SU drafted the manuscript, CL refined the draft and conceived of the research idea. All authors developed the study methods.

## 7. Acknowledgements

The authors thank Ava Kaplan, a specialist librarian at Teachers College, Columbia University, for assistance in developing the search strategy.

