## Supplementary material for "Momentary Brain Imaging Assessment of Neuroplasticity in Psychiatric Illness: Protocol for Systematic Review": Search Strategy

**Embase:**
('neuroplasticity'/exp OR 'brain plasticity':ab,ti OR 'synaptic plasticity':ab,ti)
AND
(depression/exp OR depressive disorder/exp OR mood disorder/exp OR mental health/exp OR bipolar disorder/exp OR dysthymia/exp OR cyclothymia/exp OR anxiety/exp OR panic disorder/exp OR agoraphobia/exp OR phobia'/exp OR 'anorexia nervosa'/exp OR 'bulimia nervosa'/exp OR 'binge eating'/exp OR 'feeding disorder'/exp OR 'eating disorder'/exp OR 'schizophrenia'/exp OR 'schizoaffective disorder'/exp OR 'psychosis'/exp OR 'psychotic disorder'/exp OR 'posttraumatic stress disorder'/exp OR 'feeding disorder'/exp OR 'obsessive compulsive disorder'/exp OR 'acute stress disorder'/exp OR 'prolonged grief disorder'/exp)
AND
('neuroimaging'/exp OR 'mri'/exp OR 'fmri'/exp OR 'pet'/exp OR 'spect'/exp OR 'ct'/exp OR 'eeg'/exp OR 'meg'/exp OR 'dti'/exp OR 'nirs'/exp OR 'tms'/exp OR 'vbm'/exp OR 'mrs'/exp)

**Pubmed:**
psychiatr* OR Any Field: mental health OR Any Field: depress* OR Any Field: mood dis* OR Any Field: bipolar OR Any Field: dysthymi* OR Any Field: cyclothymi* OR Any Field: anxi* OR Any Field: panic OR Any Field: agoraphobi* OR Any Field: phobi* OR Any Field: anorexia nervosa OR Any Field: bulimia nervosa OR Any Field: binge eating OR Any Field: feeding disorder OR Any Field: eating disorder OR Any Field: 'schizophreni* OR Any Field: schizoaffective OR Any Field: psychosis OR Any Field: psychotic disorder OR Any Field: posttraumatic stress OR Any Field: obsessive compulsive OR Any Field: acute stress OR Any Field: prolonged grief AND (neuroimaging OR MRI OR fMRI OR magnetic resonance OR PET OR positron emission OR SPECT OR Single-photon emission computed tomography OR CT computed tomography OR EEG OR electroencephalograph* OR MEG OR magnetoencephalograph* OR DTI OR diffusion OR NIRS OR fNIRS OR near-infrared radiation OR transcranial magnetic OR TMS OR spectroscopy) AND (neuroplast* OR brain plast* OR synaptic plast* OR glia plast* OR paired associative)

**PsychINFO:**
(Any Field: neuroimaging OR Any Field: MRI OR Any Field: fMRI OR Any Field: magnetic resonance OR Any Field: PET OR Any Field: positron emission OR Any Field: SPECT OR Any Field: Single-photon emission computed tomography OR Any Field: CT computed tomography OR Any Field: EEG OR Any Field: electroencephalograph* OR Any Field: MEG OR Any Field: magnetoencephalograph* OR Any Field: DTI OR Any Field: diffusion OR Any Field: NIRS OR Any Field: fNIRS OR Any Field: near-infrared radiation OR Any Field: transcranial magnetic OR Any Field: TMS OR Any Field: spectroscopy) AND (Any Field: neuroplast* OR Any Field: brain plast* OR Any Field: synaptic plast* OR Any Field: glia plast* OR Any Field: paired associative) AND (Any Field: psychiatr* OR Any Field: Any Field: mental health OR Any Field: Any Field: depress* OR Any Field: Any Field: mood dis* OR Any Field: Any Field: bipolar OR Any Field: Any Field: dysthymi* OR Any Field: Any Field: cyclothymi* OR Any Field: Any Field: anxi* OR Any Field: Any Field: panic OR Any Field: Any Field: agoraphobi* OR Any Field: Any Field:

phobi* OR Any Field: Any Field: anorexia nervosa OR Any Field: Any Field: bulimia nervosa OR Any Field: Any Field: binge eating OR Any Field: Any Field: feeding disorder OR Any Field: Any Field: eating disorder OR Any Field: Any Field: 'schizophreni* OR Any Field: Any Field: schizoaffective OR Any Field: Any Field: psychosis OR Any Field: Any Field: psychotic disorder OR Any Field: Any Field: posttraumatic stress OR Any Field: Any Field: obsessive compulsive OR Any Field: Any Field: acute stress OR Any Field: Any Field: prolonged grief)

**Web of Science:**
(neuroplast* OR brain plast* OR synaptic plast* OR glia plast* OR paired associative [all fields]) AND (psychiatr* OR mental health OR depress* OR mood dis* OR bipolar OR dysthymi* OR cyclothymi* OR anxi* OR panic OR agoraphobi* OR phobi* OR anorexia nervosa OR bulimia nervosa OR binge eating OR feeding disorder OR eating disorder OR 'schizophreni* OR schizoaffective OR psychosis OR psychotic disorder OR posttraumatic stress OR obsessive compulsive OR acute stress OR prolonged grief [all fields]) AND (neuroimag* OR MRI OR fMRI OR PET OR SPECT OR magnetic resonance OR positon emission OR spectroscop* [all fields])
